# Self-management of multimorbidity in sub-Saharan Africa: a systematic review and meta-synthesis with focus on diabetes, hypertension, chronic kidney disease and HIV infection

**DOI:** 10.1101/2024.09.27.24314469

**Authors:** Sangwani Nkhana Salimu, Melissa Taylor, Stephen A Spencer, Deborah Nyirenda, Nicola Desmond, Ben Morton

**Affiliations:** Department of Clinical Sciences, Liverpool School of Tropical Medicine, Liverpool, UK; Malawi-Liverpool-Wellcome Programme, Blantyre, Malawi; Department of International Public Health, Liverpool School of Tropical Medicine, Liverpool, UK

## Abstract

**Background:** The increasing prevalence of multimorbidity in sub-Saharan Africa (SSA) is an urgent concern for health systems delivery. It is uncertain how best to promote self-management approaches or the actions that patients and carers take to maintain physical and mental health, in this context. This review aims to identify, critically appraise, and synthesize qualitative evidence that describes self-management of multimorbidity among patients and their carers in SSA.

**Methods:** We systematically searched PubMed, MEDLINE, CINAHL Global Health, Google Scholar and grey literature for studies on self-management of multimorbidity or common individual chronic diseases (HIV, diabetes, hypertension or chronic kidney disease) in SSA published between 1 January 2000 and 28 to 15^th^ September 2024. Using qualitative meta-synthesis techniques to formulate the questions and synthesize findings. We used a 10-point Critical Appraisal Skills Program (CASP)-Qualitative-Checklist to assess the quality of the studies and NVivo 12 software to facilitate a thematic analysis approach.

**Results:** We screened 2010 articles for inclusion and 20 studies met inclusion criteria. We identified themes related to medical, diet, emotional and physical self-management activities. Patients negotiate self-management based on immediacy of needs and available family support. Patients are motivated to apply biomedical management but are limited by factors such as drug stock-outs and out-of-pocket expenditure. Limited knowledge and low self-efficacy toward self-management of multimorbidity impact decision making and problem solving. We found that diabetes mellitus presents the biggest stressor in terms of burden of treatment; temporal discontinuation of medications is more prevalent amongst patients with hypertension; and patients with multimorbidity are frequently hypervigilant about their health, more likely to suffer from stress and to seek healthcare.

**Conclusions:** We found that there is relative lack of data on self-management of multimorbidity among patients and their carers in SSA. Where data exists, we observed significant health literacy gaps, low health literacy support and limited self-efficacy as barriers to implementation of self-management. Context sensitive programmes are required to improve health literacy to increase patient autonomy and their toolkit of options to manage chronic disease.

## Introduction

Multimorbidity, the presence of two or more long-term conditions (LTCs), is an immediate and increasing health challenge in low- and middle-income countries (LMICs) (1-3). There are concerns, however, that this definition does not address the person-centered needs of multimorbidity related to the complex relationships between patient or carer preferences, priorities, and social context (2, 4, 5). Despite increasing prevalence of multimorbidity, data on patient and carer health literacy, skills and engagement are limited in sub-Saharan Africa (SSA) (3, 6). Self-management refers to the actions individuals and carers take for themselves, their families and others to maintain good physical and mental health; meet social and psychological needs; and prevent illness deterioration (7-9). This article aims to evaluate available data on the implementation of self-management strategies for multimorbidity in SSA.

Effective self-management requires partnership with healthcare providers to support patients and carers to increase health literacy, skills and confidence (7, 8, 10, 11). There are notable differences in how multimorbidity presents in SSA compared to high-income countries (HIC); people in SSA more frequently suffer from co-existing chronic non-communicable diseases (NCD) and communicable diseases such as HIV-infection (3, 12, 13). This fact, combined with limited access to care and poorly developed pathways for the prevention and management of chronic conditions (14), pose particular challenges for patients. People living with multimorbidity (PLWMM) must constantly negotiate decisions related to their illnesses and interactions with healthcare (15-17). Individuals self-reflect and self-negotiate on pivotal occurrences related to their conditions; when/where to seek care; and what disease and/or symptoms to prioritize (18, 19). These occurrences function as critical cues or indicators that may lead to significant changes in an individual’s approach to self-care, serving as signals that prompt a reassessment and potential refinement of their management strategies. The inter-relationship and recognition of chronic and acute symptoms is a particular challenge in this respect (18, 19). Limited access to healthcare and high out-of-pocket expenditure exacerbates the challenges for PLWMM in SSA (16, 20-22). Furthermore, limited or poor-quality interactions with healthcare providers may also present missed opportunities to improve health literacy and self-management (23).

Our aim was to explore existing evidence on the implementation of self-management strategies for PLWMM, focusing on four common disease combinations (HIV, hypertension, diabetes and chronic kidney disease [CKD]) in SSA (6, 24). We conducted a systematic literature search and meta-synthesis of primary qualitative papers related to patients living with two or more of these diseases. Lessons drawn from the review will help inform the development of improved patient-centred self-management strategies for PLWMM.

## Methods

### Design

We conducted a qualitative meta-synthesis focused on patient and carer experiences of living with multimorbidity, examining current and potential self-management approaches in SSA. The systematic review protocol was registered in the International Prospective Register of Systematic Reviews (PROSPERO) database (ID: CRD42021262708). We followed methodological and reporting procedures for qualitative meta-synthesis as outlined by Sandelowsi and Barroso (2007) (25).

### Search methods

We conducted a systematic literature search within Medline, CINAHL, and Global Health (25) databases. Our detailed search terms and strategy are described in Box 1.

### Selection of studies

We included: (1) studies that employed a qualitative, or mixed methodology; (2) explored self-management of multimorbidity among adults (≥18 years old) and/or their carers living with HIV, CKD, diabetes or hypertension or any combination of these diseases; (3) published between January 2000 and September 2024; and (4) conducted in sub-Saharan Africa. We excluded (1) abstracts and conference proceedings that were not peer reviewed; (2) studies published in a language other than English; (3) systematic or literature reviews; and (4) studies focused on palliative procedures.

A minimum of two people (SNS, SAS or MT) independently screened titles, abstracts, and full text in turn for manuscript inclusion. Subsequently, any discrepancies were discussed until consensus was reached (25). We also screened reference lists of papers included in the article. Eligible papers are summarized in Table 2.

### Quality appraisal

We appraised the quality of eligible papers using the 10-item Critical Appraisal Skills Programme (CASP) checklist for systematic appraisal of qualitative research (26). SNS and SAS appraised papers independently; discussed discrepancies until they reached consensus or referred to MT for arbitration. We assessed 10 areas: aim, methodology, research design, ethical considerations, research self-scrutiny, recruitment strategy, data-collection, data-analysis, findings, and study relevance using a 3-point scale with range 0-1 (1, Yes; 0, No; 0.5, Unclear) for each area. We assigned scores as follows: a weak score, (0) to articles that offered little to no justification for a particular issue; a moderate score, (0.5), to articles that addressed the issue but did not fully elaborate (e.g., the justification for using a self-management strategy was presented but the context itself was not explained) and a strong score, (1), to articles that extensively justified and explained the issue at hand. Studies that had a combined score across all areas between 9-10 were designated as high quality. Fourteen studies described clear aims, utilized appropriate methods, and offered meaningful interpretations indicating strong methodological rigor and were adjudged to be of high quality (25). See Box 3 for critical appraisal results.

### Analysis

Qualitative data synthesis was conducted using a thematic approach informed by Braun and Clarke, 2006 (27). The first author (SNS) familiarized herself with all the papers through reading and re-reading. Data analysis involved extraction of raw data including quotes as first order constructs and author interpretations as second-order constructs. SNS then generated third-order constructs as themes that represented synthesized interpretations of the data. To increase validity of the synthesis, first order constructs were conceptualized as key concepts and we then identified quotes that best represented each second-order construct. Additional abstraction led to development of third-order constructs (new themes), which were discussed amongst the full team (ND, DN and BM). In line with established practice, where necessary, we overrode primary researchers’ interpretations in favor of direct participants accounts (28). We explored codes and examined their relationship to each other to identify themes. Data synthesis was iterative to enhance depth of findings through adding, removing or merging codes as well as re-analyzing the data upon the emergence of new themes.

## Results

### Characteristics of included studies

We screened 2010 papers in the title phase; of these, 62 underwent abstract screening and 20 studies fulfilled criteria for inclusion (Figure 1: PRISMA flow chart). Studies reported findings from seven countries: six in South Africa (23, 29-33); five in Malawi (15, 34-37); three in Ghana (38-40) and Liberia (41-43); two from Nigeria (41, 44) and one each from Senegal (45), and Kenya (46). Four papers purposively recruited PLWMM (15, 23, 34, 47) while twelve studies addressed diabetes (29, 31, 32, 35, 37-39, 42-46). Additionally, two studies covered hypertension (30, 41) and another two covered HIV (33, 34). The methods used included semi-structured in-depth interviews (IDIs) (n = 10) (15, 29, 33, 34, 37, 38, 40, 41, 44, 45); IDIs and observations (n= 3) (23, 31, 46); FGDs (n = 1) (35); IDIs and FGDs (n=3) (30, 32, 47); IDIs, FGDs and observations (n = 1) (39); and photovoice (n=2) (42, 43). Two studies employed mixed methods (44, 47). Whilst we focused on HIV, diabetes, hypertension and chronic kidney disease, eight additional chronic conditions were observed and discussed in selected studies. These included: stroke, gout, asthma, depression, sickle cell anemia, epilepsy and chronic obstructive pulmonary disease. These conditions were sometimes mentioned in combination with the primary conditions of interest. Although they are not the focus of our manuscript, their inclusion in the studies reflects their frequent co-occurrence with the primary conditions, which is crucial for understanding the complexity of managing multiple chronic diseases. This broader context helps to provide a more comprehensive view of the challenges associated with multimorbidity.

**Fig 1:**
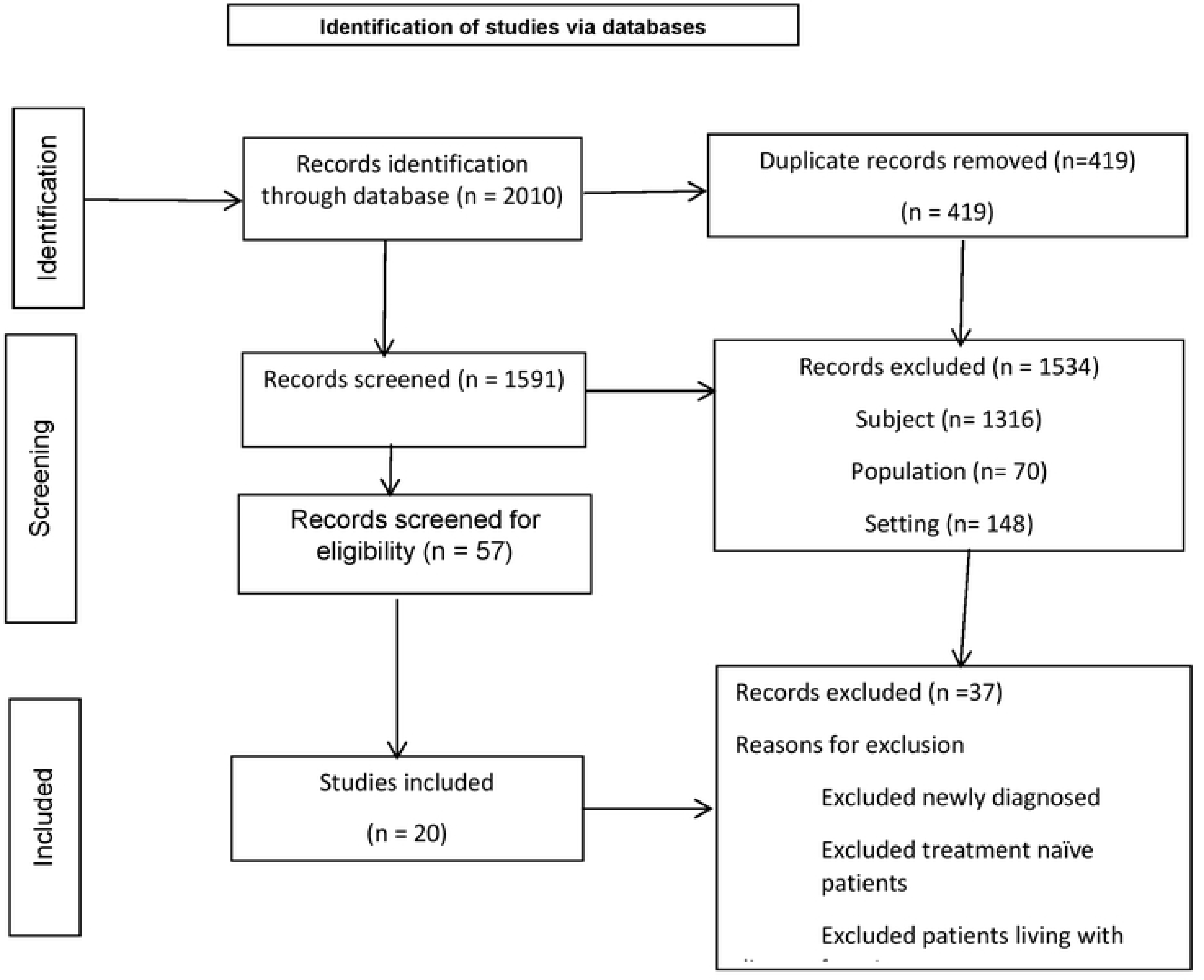
PRISMA flow chart.

### Synthesis of findings

We carried out a meta synthesis review on self-management of multimorbidity among patients living with a combination of HIV, diabetes, hypertension or chronic kidney disease. We synthesized findings under four main inductive themes namely: i) medical management; ii) diet management; iii) emotional management; and iv) physical management.

## Medical management

### Biomedical treatment is prioritized, but access is limited

The papers reviewed reported high awareness of the need for biomedical treatment among patients and carers. Commonly practiced medical management strategies include active search for medical information (15, 39); maintenance of medical appointments and drug adherence (23, 31, 32, 35, 39-41, 46-48); self-monitoring (15, 39, 40, 42, 44, 46); and avoidance of trigger factors, for example wearing tight shoes for diabetic patients (31, 36, 40). Biomedical management of chronic conditions was considered central towards optimal disease control, even more so when implemented alongside dietary and spiritual management (15, 23, 29-32, 34, 35, 39, 43, 45, 49). Patients living with diabetes fear serious consequences of hypoglycemia or hyperglycemia and claim increased treatment compliance than the other diseases (39). PLWMM are more likely to implement biomedical management to prevent secondary complications (31, 34, 39, 49).

> *“I am always afraid of low blood sugar because I have experienced it on several occasions. I go about with cube sugar in case of sudden low blood sugar. I also take cola drink whenever I have low blood sugar which I do with caution to avoid elevated blood sugar level’. (FGD 4, 48-year-old Civil servant)* (44).
>
> *“BP* (hypertension) *is dangerous because if people have not gone to the hospital, they may suffer a stroke or lose the use of some limbs or even die. Sometimes we hear of people just dying without getting sick,”* (male, 58, 13 years on ARTs, never misses ARTs-also antihypertensive).
>
> *“…I also have diabetes, and I spend most of my savings on going to the chemist to check my blood sugar level…I was admitted last month for hyperglycemic coma so the doctor said I should be checking my blood sugar every weekend…”* (Patient living with hypertension and diabetes) (41).

Patients living with singular disease experience more treatment satisfaction compared to PLWMM, reflections on treatment effectiveness relate to initiation of treatment and were directly linked to reduced distress and improved functionality (15, 34, 37, 47).

> *“Before I started taking my medication my heart could beat faster. In the case of diabetes, before I had knowledge of it I couldn’t cultivate* (farm) *because I felt very weak and I couldn’t figure it out that it was due to diabetes, but after I started taking medication now I can cultivate for long time [. . .] unlike before,”* Female, 55 diabetes and hypertension (15).
>
> *“. . .the time when I did not know of my status. . .I was very sick not expecting to be alive up to date like the way I am. I was not able to eat, to talk to anyone, or even to sit like this, no! I could only sleep and maybe do my toilet right where I was. But just after I started taking my medicine. . .I see that I am getting much better,”* IDI Female, HIV (47).
>
> *“A lot has changed in my life because at first, I did not have good health like this. My life was tough because I was suffering a lot and lost weight but when I got diagnosed and initiated on treatment I have seen great improvement in my general health and I have even gained more weight than before I started treatment,”* Man, diabetic (37).

Post initiation of treatment, most PLWMM observe and monitor how their bodies function living with disease (23, 29, 32, 39, 40, 44, 47). Patients living with HIV comorbidity are more likely to pay attention to the other disease than the HIV as many perceive their HIV to be well-managed (23, 29, 30, 32, 39, 40, 47). Again, symptom recognition related to diabetes or hypertension is a more complicated process than in HIV. This may also demonstrate that patient experiences of comorbidity may be more subjective or prolonged for diabetes, hypertension and chronic kidney disease than for HIV itself.

### Rationing drug supply as a strategy to manage access to drugs

Medication stock-outs and high costs were reported in all studies (15, 23, 29-32, 34, 35, 38-40, 44-47). Regular drug stockouts likely indicate that shortage of medication for chronic conditions in SSA is a regional challenge. Patients and their carers negotiate such challenges in multiple ways. Most studies indicate that NCD clinics are not integrated, requiring PLWMM to make multiple trips for each disease. For patients with HIV comorbidity, most papers report that patients access ART care nearby and visit a hospital further away for the other disease like diabetes or hypertension. Although ART care is generally decentralized to primary health systems, this review highlights that lack of integrated and coordinated care at tertiary facilities compounds burden of treatment through increased time and financial demands for PLWMM (35, 37, 47).

> *“I always get my ARVs from Ntabiseng clinic in Bara, and my diabetes pills from the diabetes clinic,” female living with HIV and diabetes (23)*.
>
> *“Transportation is difficult because I am not working. I attend four clinics [. . .]. Today I am here, then next month I’ll attend two other clinics.” (Person living with multimorbidity) (23)*

While challenges persist, one study reported enhanced access to information, health education and medication availability improved diabetes self-management amongst patients in Malawi (37). This study aimed to explore self-management of diabetes at a facility implementing the World Health Organization’s Package of Essential NCD Interventions for primary health care in low-resource settings (WHO PEN-Plus) (37).

We found that patients prioritize obtaining treatment despite financial concerns. Patients perceive the health outcomes of missing diabetes and hypertension treatment to outweigh the financial related costs due to transport, thus informing the decision making process (47, 50). Out-of-pocket expenditures thus indicate a strong commitment to self-management among patients and their carers. Some patients modify treatment schedule and ration drugs titrated against symptoms of disease, a practice observed particularly among hypertensive patients (34, 38, 47), who may also purchase medications from informal sources like street vendors due to lower costs (47). Some patients, however, cannot afford medications representing a significant strain on financial well-being (36, 42). This leads to delayed or absent treatment, potentially leading to disease complications, while others may turn to alternative treatment pathways, such as traditional care (23, 40, 46, 47) or apply dietary management as an alternative, rather than complimentary treatment (46). Apart from limited access to biomedical treatment, the need to access care from multiple providers is also described by patients or carers who expressed dissatisfaction with biomedical treatment; perceived that treatment was too slow; or experienced side effects (23, 40, 45, 47). This requirement to access multiple providers may be associated with decreased use of biomedical care.

> *“OK, we’ve tried the Western type of medicine for a long time …but it’s not getting better as we would want it; we want see the other aspect the traditional and the religious one, see whether it will help him to be fine, even if he will be able to stand on the other foot.” (Guardian to a diabetic single amputee) (38).”*

In some instances, patients may favor spiritual or traditional management over biomedical care but these indicate change in self-management after experiencing disease deterioration (23, 34, 39, 40, 46, 47). In such cases, biomedical treatment is sometimes sought after experiencing an acute event (39, 40), some adopting exclusive medical treatment plans to minimize future disease complications (23, 39).

Multiple medical appointments lead to increased financial costs for patients (23, 29, 30), including failure to make medical appointments for fear of confrontation by health workers for seeking care outside the allocated day (23, 29, 30). Sometimes traditional vertical models of care delivery for PLWMM results in contradictory information regarding handling of conditions and decreased confidence in health care worker capacity to address complex cases (15, 23, 29).

> *“The problem is that one doctor will tell you to do this and another asks you to do a different thing. I see different doctors for each disease. Last week, the Rheumatologist told me that my bones are getting closer to each other, they have inserted metals in my right foot. When I attended the diabetes clinic [earlier], the doctor asked me to exercise because I was adding more weight, but I can’t exercise because of the surgery they did on my leg. My ARVs have amplified my appetite. I eat a lot and I am worried about my weight too. Last time I asked the endocrinologist what else I could do to deal with my appetite, my weight gain and inability to exercise, but he just looked at me without saying a word.*” IDI, Female, 64, HIV, HTN, DM and arthritis (23).

### Mistrust presents barriers to implementation of biomedical care

Poor communication around the implementation of medical management for disease may create mistrust among patients and their carers. Misinformation or incomplete information can impact on medication adherence amongst patients or carers, presenting a significant barrier to biomedical care (31, 46). Treatment non-adherence emerges as partial, with deliberate on-and-off uptake, or complete non-uptake, with alternatives such as diet or spiritual management implemented as stand-alone forms of care (46).

> *“Why do they give me free medicines for this diabetes? They know that it will never be cured. A real medicine is never free.”* (31)
>
> *“I have purchased medicines that were fake . . . it is known that some make fake drugs so they can make lots of quick cash.”* (Diabetic individual) (46).
>
> *“When they prescribe me pharmaceutical drugs, I refuse, because I have heard that once you are used to the medicine, you have to take them every day,”* (Diabetic individual) (46).

One study reports community delivery of diabetes drugs in South Africa improved access to drugs (29). This adds on to evidence that availability of drugs closer to home improves treatment adherence (29, 37).

### Stigma associated with chronic disease

Patients with multimorbidity may experience stigma attached to individual chronic conditions such as HIV. In some cases, disclosure of conditions is secondary to maintaining social relationships especially among individuals living with HIV. Thus the value assigned to one’s social identity may negatively affect treatment adherence (15, 23, 45, 47) and potentially increase the risk of disease complications (29, 47).

> *[. . .] they advised me that I should start taking medicine. They also advised me that I should be using condoms during sexual intercourse. So, there were many things that they advised me [. . .] They advised me this, but because I went there to attend antenatal clinic and I was also feeling well, I considered that as a useless thing [. . .] I just said aaah, with the behaviour of my husband how will I initiate this? I decided that I should first deliver my baby,”* Female <50 HIV+HTN (47).

### Dietary self-management practices

Studies highlighted the importance of dietary management among PLWMM but that implementation is impacted by poverty and high cost of food (15, 31, 35, 37, 38, 41, 49); limited knowledge on portion size estimates (35, 46); and complexity of conditions and patients’ contrary cultural values (32, 35, 45, 46). Studies highlighted that preparation of separate meals for patients with chronic disease (32, 46, 47) was challenging, citing poverty and increased workload related to food preparation as the main reasons for not following dietary requirements (32, 35, 39, 44, 45, 47).

Patients and carers demonstrate of diet management are varied (41-43). Lay descriptions of recommended dietary practices include reducing the consumption of carbohydrates or sugar. However, these practices indicate faulty understanding of pertinent issues to mean only sweet foods contain sugar or starch is only present in flour. It also highlights limited comprehension of the need to balance amounts and time of consumption. Some patients and their carers suggest budgetary prioritization to optimize dietary management strategies (43).

> “*Sometimes these days I do not eat breakfast because I think after dinner I’m satisfied until lunch.”* (diabetic patient) (46) or not eating *‘starch’* but eating ‘*rice’* (diabetic patient) *(38)*.
>
> *“Oil that settles off when placed down can give you the illness […],”* (42-year-old diabetic patient) (39).
>
> *“If I want to eat the rice, my wife put the rice on a fire and put water on it, she will let the rice boil. Then she takes the starch and waste it. She put clean water on the rice again and wash it. Then she wastes it and then she let the rice steam. It shouldn’t be soft, it should be, you know, just normal rice. Then the rice does not contain no more starch,” man,* diabetic, 62, retired college graduate (43).

The inaccurate interpretations regarding what is starch or sugar, or the physical presentation of cooking oil stress that knowledge on what, how, when or how much negatively affects dietary implementation for disease. The challenge of comprehending dietary recommendations can create confusion when advice suggests moderating rather than completely eliminating certain foods (23, 34).

> *“When I go to the private clinic the doctors advise me to stop taking meat, salt, sweet beverages, and not to use cooking fat/oil, but without telling me the reasons. When I come to the public hospital the nurse explains that I should take food in moderation, so I don’t know who to believe,” Female 58, overweight, diabetic*.

Diet management relates to social factors, kinship, cultural beliefs and systems indicating an intricate relationship between cultural beliefs and what people eat (32, 35, 42, 43, 45, 46). We note that cultural identity, family influence, or community norms may significantly impact dietary practices, sometimes leading to a preference for traditional foods that do not align with medical dietary recommendations. Two studies indicate patient’ implementation of contrary practices indicative of the value placed in cultural identity (31, 45).

> *“Being African, I eat rice for lunch because it is part of my culture and Senegalese men like to eat fatty and oily “thiebou dien” every day* (Diabetic patient) *(45)*.

Desperation or defeatist attitude emerge among diabetic patients who feel hopeless (31, 45, 46) citing diabetes: *“diabetes is a slow poison”* (35, 46). Such an attitude may normalize certain behaviors contrary to recommended practices for PLWMM. Alternatively, it may indicate a lack of assertiveness or demotivation among patients, who may feel overwhelmed. This perspective can also weaken patients’ resolve to adhere to their treatment plans if they perceive their efforts as futile against the challenges posed by their condition.

> *“There are several people that I have met who do not care [about diabetes]. During social events such as wedding, prayers, or funeral, you will see them overeat . . . they tell me . . . you only die once.”* (Man, diabetic) (46).
>
> *” You cannot tell your wife that these days do not prepare this. Because you are the only one with diabetes. . .. You must be flexible . . . the issue of dietary restrictions is problematic.”* (Diabetic man) (46).

### Psychological stress among PLWMM

Patients and carers within the included studies reported that anxiety and depression are more pronounced among PLWMM than with single chronic disease (15, 23, 29, 31, 33, 34, 47). Patients with both diabetes and chronic kidney disease had particularly high levels of stress (15, 23). Studies reported that a single diagnosis of hypertension or HIV were associated with lower stress levels (47). Reported stressors included pain; deterioration of disease; onset of new illness and symptoms. These were exacerbated by poverty and limited access to biomedical treatment. For effective emotional management of chronic disease, patients must be able to risk assess; manage time; communicate; and think analytically. These processes facilitate problem solving; allocation of scarce resources; prioritization of conditions; and coordination of self-care (33-35, 47). In most cases, the dominant sources of information include family members (including those who live or had provided care to family member with a similar disease), other patients and health workers (15, 23, 39, 51). The studies demonstrate that PLWMM avoid stressors that may complicate their conditions (15, 29, 34, 35, 39).

> *“I must also keep my temper in check as I can’t afford to lose it otherwise, I risk escalating my blood sugar level and blood pressure”.* (Woman with hypertension and diabetes) (29).

Depression is cited by PLWMM as they negotiate the burden of conditions (23) or disrupted social relationships (15, 32, 47).

### Lay understanding and implementation of physical activity

PLWMM and their carers recognise the benefit of regular physical activity towards functional performance (15, 30). Some studies highlight the link between physical management and wellbeing (15, 32).

> *“I bought a testing device which I use to check myself every day. When I was employed, it was helping me because I was walking a long distance to minibus depot which was part of exercise and with the nature of my work, I was working under the sun and I was sweating, so it was also part of my physical exercise and when I test my sugar the following morning it was at a good level such as 90,”* Male <50 hypertension and diabetic (15).

For women living with multimorbidity, physical exercise was embedded within daily living activities such as domestic chores like cleaning, washing and walking to work (35, 40, 44) rather than public exercise (23, 40, 46). This integration of physical exercise into daily living activities offers a practical, accessible, and sustainable approach to improving health outcomes for PLWMM (35, 47).

Barriers to physical management for PLWMM were pain, age, lack of clarity on nature of exercise and competing priorities (15, 29, 31, 32, 39-41, 47). Anxiety, fear of exertion and motivation affect implementation of physical exercise (23, 44). Fear stems from concerns about worsening their condition, experiencing pain, or facing physical limitations, thereby creating a cycle of avoidance and inactivity that perpetuates disease complications (32).

> *“… with me I don’t walk long distances at all because of my chest, I get a tight chest when I walk a distance and I start coughing,”* Patient with hypertension (30).
>
> *‘I do try to exercise sometimes, but you know I get tired … and I will do it for a month only, then I just get tired. I am not lazy, but ….’* Female, hypertension (30).

In one study, physical management is applied as an alternative in the absence of diet management.

> ‘To deal with these challenges, Participant Y has opted to pay for a gym, which he finds to be affordable at R340 (US$22) a month, compared with buying healthy foods which he says *“are expensive.”’* Man, 67, HTN and DM (23).

### Social capital for managing multimorbidity

Social support was found to facilitate self-management in all included studies (23, 31, 32, 39, 40, 46, 47, 50). Where disclosure of condition(s) is accepted, the patient or carer leverages on social capital and gets support for financial, physical or emotional management (15, 33-35, 39). Conversely, lack of social support means patients learn to manage other people’s expectations (33, 45, 49), and sometimes, self-isolate where community expectations contradict recommended medical care (15, 44). Failure to manage external expectations sometimes leads to feelings of worthlessness among patients (15, 31). Challenges to emotional support include competing priorities among individuals who can provide support, family and social commitments and PLWMM themselves being carers to others living with chronic conditions [19, 28, 30]. The absence of emotional support negatively affects relationships (15, 29, 47). Peer support is common among patients with HIV, whether as singular or comorbid conditions (23, 29, 32, 34, 35, 39, 44, 47).

## Discussion

These results offer a comprehensive understanding of the experiences and coping strategies related to self-managing multimorbidity among patients and their carers in sub-Saharan Africa. This is important because effective self-management strategies are key to controlling chronic conditions, preventing complications, and improving quality and longevity of life. Further work is required to explore how patients and carers can be empowered to manage multimorbidity. We recommend that health literacy programmes are tailored to context to ensure that recommendations are sustainable and deliverable within the healthcare system and that advice is tailored toward social and cultural norms and practical realities.

We found high intention to use biomedical treatment among patients with multimorbidity or individual chronic diseases in SSA (15, 23, 29-32, 34, 35, 38-40, 44-47). However, multiple factors limit implementation of effective self-management strategies. In SSA, patients frequently must access multiple providers and incur out-of-pocket expenses to obtain medications, increasing treatment burden and financial strain (16, 52, 53). Financial constraints require that PLWMM often need to ration drugs or compromise dietary choices. Clearly, working toward universal healthcare coverage (UHC) would help to address some of these issues. Whilst health ministries develop and implement UHC, however, pragmatic sustainable guidance should be developed for patients to help them make optimal prioritisation decisions based on their individual circumstances. Work to improve health literacy and advocacy in patients and their carers is important to drive self-efficacy and self-management of multimorbid disease with examples in Malawi (54), Ethiopia (55), Uganda (56) and South Africa (16, 20). Building social support facilities may also help PLWMM and their carers navigate perceived barriers to self-management (57). In Kenya, for example, PLWMM who attended support groups demonstrated improved decision making and treatment compliance (58).

Our meta-synthesis shows that PLWMM and their carers apply a biosocial lens to self-management and implement strategies between conditions according to their perceived understanding of disease etiology and severity. A systematic review on determinants of HIV treatment adherence in SSA reports that adherence is linked to patient perceptions towards improved health outcomes (59). A key priority, therefore, for PLWMM in SSA would be to link management strategies with measures of disease control.

Creating these feedback loops would help to reinforce self-management decisions and priority setting for both patients and their carers. For example, temporary drug withdrawal was commonly reported among patients with hypertension. This may reflect fear of long-term treatment (58, 60) and/or communication challenges between healthcare workers and patients (60). Measures to increase patient autonomy and ownership of their hypertension together with a disease control feedback loop (e.g. blood pressure control measurements) could promote improved self-management decisions. Strategies to improve physical activity in PLWMM need to be tailored toward individuals so that they are meaningful but also relevant and sustainable. Limited knowledge and low self-efficacy are currently reported as barriers to physical exercise for PLWMM within the current literature (30, 52, 61, 62).

## Strength and limitations

To our knowledge, this study is the first qualitative meta-synthesis review to synthesize lived experiences regarding self-management of multimorbidity among patients living with a combination of diabetes, hypertension, chronic kidney disease or HIV. Our authorship group represents a multidisciplinary team including both clinical and qualitative expertise to facilitate rigorous meta-synthesis approaches and clinical relevance. The strength of our findings are limited by the relative paucity of relevant literature in this area. Exclusion of studies not published in English is a limitation and may reduce the generalisability of our findings across different linguistic and cultural contexts. We did not include quantitative data from included papers that utilised mixed methods within our qualitative synthesis. The review may therefore overlook important numerical insights that could contribute to a more comprehensive understanding of the topic.

## Conclusion

We found that PLWMM and their carers were motivated to self-manage their conditions but face multiple barriers including both financial and health systems constraints. Health systems strengthening is essential to develop more horizontal and holistic models of care delivery to reduce the need for multiple healthcare interactions and promote joined up care. Contextually sensitive measures to improve health literacy and autonomy should be prioritised as a component of this strengthening to help patients and their carers develop the tools required to manage their long-term health conditions. The development and support of community groups may bolster this process where members can support delivery of appropriate lay information and provide support to patients, reducing pressure on health systems.

## Data Availability

Thank you for your inquiry regarding the underlying data for our manuscript. We would like to clarify that our review utilized secondary data sourced from publicly available manuscripts in open-access journals. As such, we do not have any additional datasets beyond the supporting information already shared with the journal and the reviewed articles, which are all accessible online. Since we conducted a systematic review and meta-synthesis of existing literature, we do not have original data to share. We have ensured that all sources used are properly cited and available for review

## Acknowledgements

We would like to acknowledge Ms. Alison Derbyshire, the Academic Liaison & Training Specialist, Liverpool School of Tropical Medicine (LSTM) for her assistance in the search strategy. We would also like to acknowledge the authors of the original manuscripts for their dedication to this topic and for elevating the voices of marginalized people in sub-Saharan Africa.

## References

1. Price AJ, Jobe M, Sekitoleko I, Crampin AC, Prentice AM, Seeley J, et al. Epidemiology of multimorbidity in low-income countries of sub-Saharan Africa: Findings from four population cohorts. PLOS Glob Public Health. 2023;3(12):e0002677.

2. Academy of Medical Sciences. Improving the prevention and management of multimorbidity in sub-Saharan Africa. London; 2019.

3. Gouda HN, Charlson F, Sorsdahl K, Ahmadzada S, Ferrari AJ, Erskine H, et al. Burden of non-communicable diseases in sub-Saharan Africa, 1990–2017: results from the Global Burden of Disease Study 2017. The Lancet Global Health. 2019;7(10):e1375-e87.

4. Dixon J, Morton B, Nkhata MJ, Silman A, Simiyu IG, Spencer SA, et al. Interdisciplinary perspectives on multimorbidity in Africa: developing an expanded conceptual model. medRxiv. 2023:2023.09.19.23295816.

5. Banda G, Bosire E, Bunn C, Chandler C, Chikumbu E, Chiwanda J, et al. Multimorbidity research in Sub-Saharan Africa: Proceedings of an interdisciplinary workshop [version 1; peer review: 1 approved with reservations]. Wellcome Open Research. 2023;8(110).

6. Spencer SA, Rylance J, Quint JK, Gordon SB, Dark P, Morton B. Use of hospital services by patients with chronic conditions in sub-Saharan Africa: a systematic review and meta-analysis. Bull World Health Organ. 2023;101(9):558–70g.

7. Lorig K. Self-Management of Chronic Illness: A Model for the Future. Generations: Journal of the American Society on Aging. 1993;17(3):11–4.

8. Schulman-Green D, Jaser S, Martin F, Alonzo A, Grey M, McCorkle R, et al. Processes of self-management in chronic illness. J Nurs Scholarsh. 2012;44(2):136–44.

9. Aantjes CJ, Ramerman L, Bunders JFG. A systematic review of the literature on self-management interventions and discussion of their potential relevance for people living with HIV in sub-Saharan Africa. Patient Education & Counseling. 2014;95(2):185–200.

10. World Health Organization. Multimorbidity: Technical Series on Safer Primary Care. Geneva: World Health Organization; 2016. Report No.: Licence: CC BY-NC-SA 3.0 IGO.

11. Grady PA, Gough LL. Self-management: a comprehensive approach to management of chronic conditions. Am J Public Health. 2014;104(8):e25–31.

12. Asogwa OA, Boateng D, Marzà-Florensa A, Peters S, Levitt N, van Olmen J, et al. Multimorbidity of non-communicable diseases in low-income and middle-income countries: a systematic review and meta-analysis. BMJ Open. 2022;12(1):e049133.

13. Academy of Medical Sciences. Multimorbidity: a priority for global health research. Google scholar. 2019.

14. Yadav UN, Lloyd J, Hosseinzadeh H, Baral KP, Bhatta N, Harris MF. Self-management practice, associated factors and its relationship with health literacy and patient activation among multi-morbid COPD patients from rural Nepal. BMC Public Health. 2020;20(1):300.

15. Chikumbu EF, Bunn C, Kasenda S, Dube A, Phiri-Makwakwa E, Jani BD, et al. Experiences of multimorbidity in urban and rural Malawi: an interview study of burdens of treatment and lack of treatment. PLoS Global Public Health. 2022;2(3).

16. van Pinxteren M, Mbokazi N, Murphy K, Mair FS, May C, Levitt N. The impact of persistent precarity on patients’ capacity to manage their treatment burden: A comparative qualitative study between urban and rural patients with multimorbidity in South Africa. Frontiers in Medicine. 2023;10.

17. Tran PB, Ali A, Ayesha R, Boehnke JR, Ddungu C, Lall D, et al. An interpretative phenomenological analysis of the lived experience of people with multimorbidity in low- and middle-income countries. BMJ Global Health. 2024;9(1):e013606.

18. Shippee ND, Shah ND, May CR, Mair FS, Montori VM. Cumulative complexity: a functional, patient-centered model of patient complexity can improve research and practice. J Clin Epidemiol. 2012;65(10):1041–51.

19. Ridgeway JL, Egginton JS, Tiedje K, Linzer M, Boehm D, Poplau S, et al. Factors that lessen the burden of treatment in complex patients with chronic conditions: a qualitative study. Patient Prefer Adherence. 2014;8:339–51.

20. Roomaney RA, van Wyk B, Pillay-van Wyk V. Multimorbidity in South Africa: Is the health system ready? J Multimorb Comorb. 2023;13:26335565231182483.

21. Coventry PA, Small N, Panagioti M, Adeyemi I, Bee P. Living with complexity; marshalling resources: a systematic review and qualitative meta-synthesis of lived experience of mental and physical multimorbidity. BMC Family Practice. 2015;16(1):171.

22. Younas A, Inayat S. Alleviating suffering of individuals with multimorbidity and complex needs: A descriptive qualitative study. Nursing Ethics.0(0):09697330231191280.

23. Bosire EN. Patients’ Experiences of Comorbid HIV/AIDS and Diabetes Care and Management in Soweto, South Africa. Qualitative health research. 2021;31(2):373–84.

24. Gouda HN, Charlson F, Sorsdahl K, Ahmadzada S, Ferrari AJ, Erskine H, et al. Burden of non-communicable diseases in sub-Saharan Africa, 1990–2017: results from the Global Burden of Disease Study 2017. The Lancet Global Health. 2019;7(10):e1375-e87.

25. Sandelowski M, Barroso J, Voils CI. Using qualitative metasummary to synthesize qualitative and quantitative descriptive findings. Res Nurs Health. 2007;30(1):99–111.

26. Critical Appraisal Skills Programme (CASP). Critical Appraisal Skills Programme (CASP). 2013. Qualitative research checklist URL: http://media.wix.com/ugd/. 2013.

27. Braun V, Clarke V. Using thematic analysis in psychology. Qualitative Research in Psychology. 2006;3(2):77–101.

28. Ludvigsen M, Hall E, Meyer G, Fegran L, Aagaard H, Uhrenfeldt L. Using Sandelowski and Barroso’s Meta-Synthesis Method in Advancing Qualitative Evidence 2016.

29. Mendenhall E, Norris SA. Diabetes care among urban women in Soweto, South Africa: a qualitative study. BMC public health. 2015;15:1300.

30. Magobe NBD, Poggenpoel M, Myburgh C. Experiences of patients with hypertension at primary health care in facilitating own lifestyle change of regular physical exercise. Curationis. 2017;40(1):e1–e8.

31. Matwa P, Chabeli MM, Muller M, Levitt NS. Experiences and guidelines for footcare practices of patients with diabetes mellitus. Curationis. 2003;26(1):11–21.

32. Steyl T, Phillips J. Management of type 2 diabetes mellitus: adherence challenges in environments of low socio-economic status. African journal of primary health care & family medicine. 2014;6(1):E1–E7.

33. Tyabazeka S, Phiri W, Marie Modeste RR. HIV self-management perceptions and experiences of students at one university in South Africa. Curationis. 2024;47(1):e1-e10.

34. Moucheraud C, Phiri K, Hoffman RM. Health behaviours and beliefs among Malawian adults taking antihypertensive medication and antiretroviral therapy: A qualitative study. Global public health. 2022;17(5):688–99.

35. Mphwanthe G, Carolan M, Earnesty D, Weatherspoon L. Perceived barriers and facilitators to diet and physical activity among adults diagnosed with type 2 diabetes in Malawi. Global public health. 2021;16(3):469–84.

36. Angwenyi V, Aantjes C, Bunders-Aelen J, Lazarus JV, Criel B. Patient–provider perspectives on self-management support and patient empowerment in chronic care: A mixed-methods study in a rural sub-Saharan setting. Journal of Advanced Nursing (John Wiley & Sons, Inc). 2019;75(11):2980–94.

37. Drown L, Adler AJ, Schwartz LN, Sichali J, Valeta F, Boudreaux C, et al. Living with type 1 diabetes in Neno, Malawi: a qualitative study of self-management and experiences in care. BMC health services research. 2023;23(1):595.

38. Aikins AdG, Awuah RB, Pera TA, Mendez M, Ogedegbe G. Explanatory models of diabetes in urban poor communities in Accra, Ghana. Ethnicity & Health. 2015;20(4):391–408.

39. de-Graft Aikins A. Healer shopping in Africa: new evidence from rural-urban qualitative study of Ghanaian diabetes experiences. BMJ (Clinical research ed). 2005;331(7519):737.

40. Amu H, Darteh EKM, Tarkang EE, Kumi-Kyereme A. Management of chronic non-communicable diseases in Ghana: a qualitative study using the chronic care model. BMC public health. 2021;21(1):1120.

41. Ukoha-Kalu BO, Adibe MO, Ukwe CV. A qualitative study of patients’ and carers’ perspectives on factors influencing access to hypertension care and compliance with treatment in Nigeria. Journal of hypertension. 2023;41(5):845–51.

42. Bleah P, Wilson R, Macdonald D, Camargo-Plazas P. ’The solution is we need to have a centre’: a study on diabetes in Liberia. Health promotion international. 2023;38(5).

43. Bleah P, Wilson R, Macdonald D, Camargo Plazas P. “When I Don’t Have Money, I Don’t Eat”: A Critical Hermeneutic Study of Diabetes in Liberia2023.

44. Okurumeh AI, Akpor OA, Okeya OE, Akpor OB. Type 2 diabetes mellitus patients’ lived experience at a tertiary hospital in Ekiti State, Nigeria. Scientific reports. 2022;12(1):8481.

45. BeLue R, Diaw M, Ndao F, Okoror T, Degboe A, Abiero B. A cultural lens to understanding daily experiences with type 2 diabetes self-management among clinic patients in M’bour, Senegal. International quarterly of community health education. 2012;33(4):329–47.

46. Abdulrehman MS, Woith W, Jenkins S, Kossman S, Hunter GL. Exploring Cultural Influences of Self-Management of Diabetes in Coastal Kenya: An Ethnography. Global qualitative nursing research. 2016;3:2333393616641825.

47. Angwenyi V, Aantjes C, Kajumi M, De Man J, Criel B, Bunders-Aelen J. Patients experiences of self-management and strategies for dealing with chronic conditions in rural Malawi. PLoS ONE. 2018;13(7):1–17.

48. Mphwanthe G, Carolan MT, Earnesty D, Weatherspoon LJ. Perceived barriers and facilitators to diet and physical activity among adults diagnosed with type 2 diabetes in Malawi. Global Public Health. 2020;16:469–84.

49. Angwenyi V, Bunders-Aelen J, Criel B, Lazarus JV, Aantjes C. An evaluation of self-management outcomes among chronic care patients in community home-based care programmes in rural Malawi: A 12-month follow-up study. Health & Social Care in the Community. 2021;29(2):353–68.

50. Mphande M, Campbell P, Hoffman RM, Phiri K, Nyirenda M, Gupta SK, et al. Barriers and facilitators to facility HIV self-testing in outpatient settings in Malawi: a qualitative study. BMC Public Health. 2021;21(2200).

51. Amu H, Brinsley TY, Kwafo FO, Amu S, Bain LE. Improving investment in chronic disease care in Sub-Saharan Africa is crucial for the achievement of SDG 3.4: application of the chronic care model. Archives of Public Health. 2023;81(1):169.

52. Murphy K, Chuma T, Mathews C, Steyn K, Levitt N. A qualitative study of the experiences of care and motivation for effective self-management among diabetic and hypertensive patients attending public sector primary health care services in South Africa. BMC health services research. 2015;15:303.

53. Areri HA, Marshall A, Harvey G. Interventions to improve self-management of adults living with HIV on Antiretroviral Therapy: A systematic review. PloS one. 2020;15(5):e0232709.

54. Kwanjo Banda C, Gombachika B, Nyirenda M, Muula A. Self-management and its associated factors among people living with diabetes in Blantyre, Malawi: a cross-sectional study [version 2; peer review: 2 approved with reservations]. Open Research Africa. 2021;2(161).

55. Ferrari G, Ngoga G, Manzi A, Gomber A. Peer Support in the Management of Diabetes to Improve Cardiovascular Disease Outcomes in Low- and Middle-Income Countries (LMICs). Global Heart. 2023.

56. Tusubira AK, Nalwadda CK, Akiteng AR, Hsieh E, Ngaruiya C, Rabin TL, et al. Social Support for Self-Care: Patient Strategies for Managing Diabetes and Hypertension in Rural Uganda. Annals of Global Health. 2021.

57. Adeniyi A, Idowu O, Ogwumike O, Adeniyi C. Comparative influence of self-efficacy, social support and perceived barriers on low physical activity development in patients with type 2 diabetes, hypertension or stroke. Ethiop J Health Sci. 2012;22(2):113–9.

58. Otieno P, Agyemang C, Wilunda C, Sanya RE, Iddi S, Wami W, et al. Effect of Patient Support Groups for Hypertension on Blood Pressure among Patients with and Without Multimorbidity: Findings from a Cohort Study of Patients on a Home-Based Self-Management Program in Kenya. Global Heart. 2023.

59. Heestermans T, Browne JL, Aitken SC, Vervoort SC, Klipstein-Grobusch K. Determinants of adherence to antiretroviral therapy among HIV-positive adults in sub-Saharan Africa: a systematic review. BMJ Global Health. 2016;1(4):e000125.

60. Macquart de Terline D, Kane A, Kramoh KE, Ali Toure I, Mipinda JB, Diop IB, et al. Factors associated with poor adherence to medication among hypertensive patients in twelve low and middle income Sub-Saharan countries. PLOS ONE. 2019;14(7):e0219266.

61. Modeste RRM, Majeke SJ. Self-care symptom-management strategies amongst women living with HIV/AIDS in an urban area in KwaZulu-Natal. Health SA Gesondheid (Online). 2010;15:1–8.

62. Schulman-Green D, Jaser SS, Park C, Whittemore R. A metasynthesis of factors affecting self-management of chronic illness. J Adv Nurs. 2016;72(7):1469–89.

